# Improving COVID-19 vaccination centre operation through computer modelling and simulation

**DOI:** 10.1101/2021.03.24.21253517

**Authors:** RM Wood, SJ Moss, BJ Murch, C Davies, C Vasilakis

**Affiliations:** Modelling and Analytics; UK National Health Service (BNSSG CCG); Centre for Healthcare Improvement and Innovation (CHI^2^); School of Management; University of Bath

**Keywords:** COVID-19, Coronavirus, Mass Vaccination, Vaccination Centre, Operational Research

## Abstract

Mass vaccination is widely considered to offer the only route out of the COVID-19 pandemic in a way that social restrictions can be eased and economic activity can safely resume without compromising healthcare services. For many countries, dedicated vaccination centres are a key part of that effort. However, with no directly comparable historical experience there is little information to guide the operational configuration and management of these sites. In addressing this gap in knowledge, the objective of this study is to demonstrate the value of computer modelling. This is achieved through providing an account of its use in supporting management considerations and decisions at two major vaccination centres, at an early stage of the UK’s mass vaccination effort. We report on how modelling insight has influenced the initial setup of one site, including quantification of daily booking numbers. For the same site, we reveal how analysis has informed a significant operational shift in combining two key activities on the vaccination pathway into one. Finally, we describe how, at a second site, modelling has been used to examine pathway stability, in terms of resilience to unforeseen ‘shocks’ such as delayed arrivals and staff unavailability. Alongside the open-source simulation software, the modelling insights reported here can support managers to better plan and improve the operation of COVID-19 vaccination centres.

## 1. Introduction

On 9 November 2020 it was announced that the mRNA-based Pfizer-BioNTech vaccine was at least 90% effective in preventing COVID-19 disease [1]. Two weeks later, news broke of successful Phase 3 trials involving the Oxford-AstraZeneca vaccine – an adenovirus-based alternative of lesser cost and not requiring extreme storage temperature [2]. The UK regulator approved the Pfizer-BioNTech (PZ) vaccine on 2 December 2020 [3], with the first dose administered shortly after on 8 December 2020 [4]. Later that month the Oxford-AstraZeneca (AZ) vaccine was granted approval [5], opening up the possibility of mass vaccination across the UK [6].

Vaccination of sufficiently large proportions of the population is integral to reducing the various non-pharmaceutical interventions imposed over the course of the pandemic [7,8]. In turn, this may allow for all sectors of the economy to be safely reopened, quality of life to improve, and healthcare services to resume their normal operation – including addressing the many unmet demands sustained during the pandemic [9].

Alongside acute hospitals, pharmacies and general practice (family doctors), a key setting for mass vaccine administration is the dedicated vaccination centre. Often located in sports and leisure facilities, these are designed to provide an efficient high-throughput service for advance-booked individuals requiring vaccination. In the UK, the population is vaccinated in order of priority group, based mainly on age but also accounting for underlying health conditions and employment as a health or care worker [10].

For individuals attending a UK COVID-19 vaccination centre, there are a number of key activities that may be expected. Following arrival, the individual would register and confirm their details and booking. They would then be clinically assessed to ensure suitability for the vaccine. Then they would be vaccinated and – possibly following some observation period, as required for the PZ vaccine – they would exit the site. From the service provision perspective, there activities were predominantly organised along two pathways (Figure 1); the essential difference being that *Clinical Assessment* and *Vaccination* were either performed as separate activities by separate teams or were performed conjointly by the same team (and thus with no requirement for the vaccinee to queue in between).

**Figure 1.**
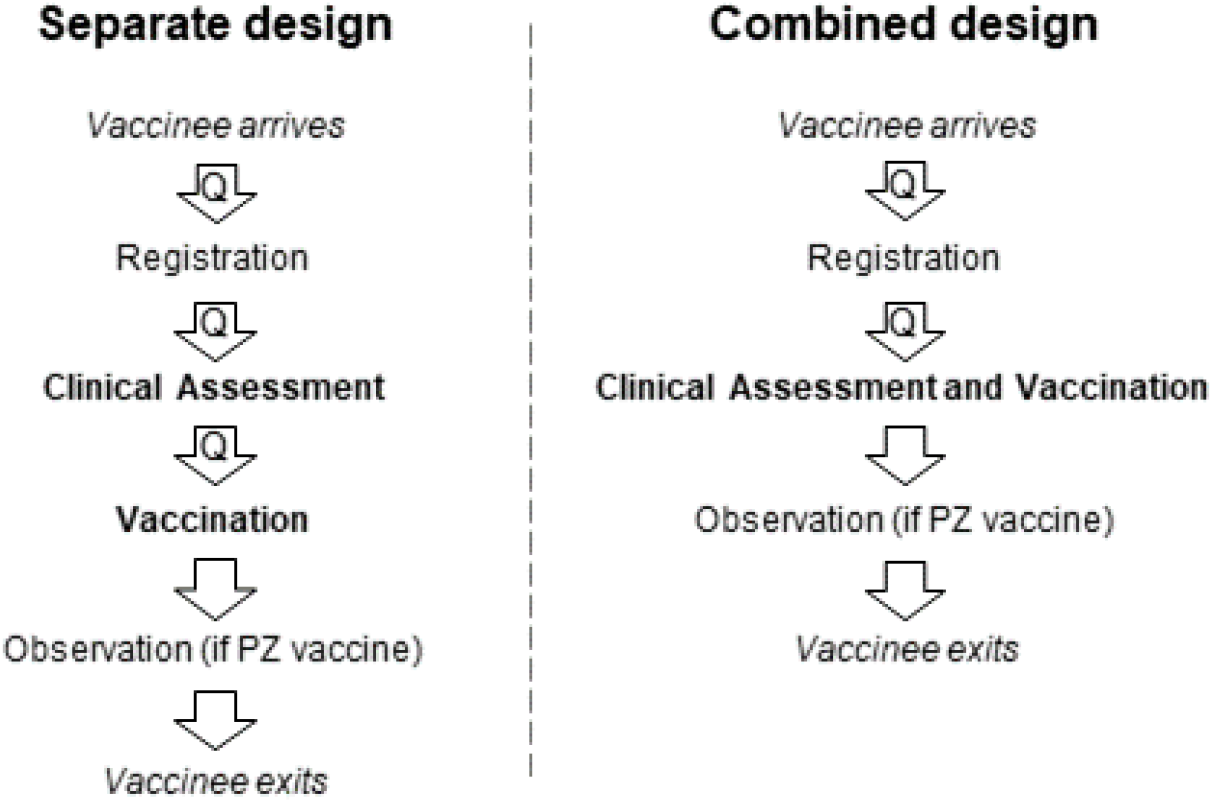
Outline of key activities along the vaccination centre pathway for both the ‘separate’ and ‘combined’ design options predominantly in use at an early stage of the mass vaccination effort in the UK. Transitions between activities where the vaccinee may be required to queue are denoted by ‘Q’.

There are a number of operational considerations in effectively configuring a vaccination centre. First, there is the number of vaccinees to book in for each operating period. Ultimately the aim is to maximise throughput, but this must be done safely. If throughput is too high then unsustainable queues will form, compromising social distancing and impairing patient experience (important for ensuring a repeat visit for any additional dose). On the other hand, if throughput is too low, then this would lead to an uneconomic use of available resources. Additional considerations relate to the optimal allocation of activity-level resources to ensure balanced server utilisation and the incorporation of sufficient ‘slack’ in pathway capacity to ensure any ‘shocks’ can be readily absorbed (such as staff sickness or a number of vaccinees arriving all at once).

There is, however, very little information and learned experience to guide managers through these considerations. Events of such magnitude have simply not occurred in recent times and so, beyond the limited number of national and regional level emergency preparedness roles, there is little existing knowledge and expertise within the local frontline entities tasked with setting up the vaccination centres. Clearly, it is very early for empirical studies on the operation of COVID-19 vaccination centres to have emerged in the peer review literature. One recently published study has used discrete-event and agent-based simulation techniques to model the operation of a drive-through mass vaccination clinic, but this has not been informed by empirical data and the tool is not open-source or freely available [11].

Previous pandemics such as H1N1 (2009-2010) as well as the general necessity for adequate emergency preparedness have provided opportunities for comparable studies to be conducted in the past. Many of these used mathematical modelling and simulation to investigate the effectiveness of different types of vaccination programmes and were conducted at a population rather than a vaccination centre level [12,13]. There are a number of simulation studies at vaccination centre/clinic level including generic frameworks that can be applied in different contexts [14,15], including antibiotic distribution centres [16], and for infections with influenza /H1N1/pneumococcal [17-22] and smallpox [23]. Almost all of these studies use the discrete-event approach to stochastic simulation, given its capacity of capturing the modelling requirements of service systems with individual entities (such as patients) which flow through a care pathway, competing for resources such as appointment slots [24].

The study reported here is, to the best of our knowledge, the first to demonstrate the value of computer modelling in supporting operational planning at COVID-19 vaccination centres. This is approached by providing an account of how modelling was used to inform the configuration of two major regional vaccination centres in South West England. Modelling of vaccination centre activities (Figure 1) is performed through a discrete event simulation approach presented in Section 2. A description of how this was used to set daily bookings and inform site design at Vaccination Centre A is provided in Section 3.1. Modelling to support the management consideration of shifting from the initially-used ‘separate’ pathway design to a ‘combined’ one is covered in Section 3.2. Vaccination Centre B – an up-and-running site operating under the ‘combined’ design – is introduced in Section 3.3, with modelled throughput compared against that of Vaccination Centre A. Finally, in Section 3.4, the operational resilience of Vaccination Centre B is tested through simulating a number of potential ‘shocks’ to the pathway. A discussion on limitations, extensibility and further applied research opportunities is presented in Section 4.

## 2. Model

### 2.1 Requirements

In representing COVID-19 vaccination centre dynamics, any solution should concern a network of multi-server service points describing the various sequential activities outlined in Figure 1. Arrivals should be assumed independent of one another, activity-level service time should be approximated by the appropriate statistical distributions, and vaccinees should be served in the order of arrival if queued due to unavailable service point capacity. While the physical dimensions of any site may necessitate a formal limit on the capacity of the various queues, it is unrealistic to account for such constraints within the model since this would assume that when such limits are breached then vaccinees will block upstream service channels. In practice, and as informed by observing a live exercise prior to the opening of either vaccination centre (more information in Section 3.1), vaccinees will continue to join the queue, albeit in a way that may compromise social distancing. In Kendall’s notation [25], a solution is therefore required to the queueing system specified by a network of of *M*| *G*| *c*| ∞| *FIFO* queues in series. That is, Markovian arrivals (*M*), general service times (*G*), a finite number of servers (*c*), an infinite queueing capacity (∞), and a first-in first-out queue discipline (*FIFO*).

Given that vaccination centres were, at least initially, expected to operate for a fixed period within each day (i.e. not continuously) the model should capture transient behaviour across the given operating period. To be useful for management, the modelled outputs should present, over time, the number of vaccinees within each activity and (where relevant) the number of vaccinees queueing for each activity. Agreed as key measures of interest by site managers, the former provides an assessment of resource utilisation along the vaccination pathway, including whether activity-level resources are evenly used, while the latter allows for consideration of safety and patient experience. For such measures, confidence bands are required to facilitate an assessment of variability throughout the operating period. Site managers had indicated that these could be used to gauge the suitability of any configuration through some notion of risk appetite, e.g. a configuration would be inappropriate if the upper 95% confidence band on queue size exceeds the dedicated queue capacity during the operating period.

### 2.2 Implementation

Computer simulation was used to model the vaccination centre pathway. An analytical solution was not attempted due to limitations on versatility (e.g. the ability to introduce temporal dependence on the arrival rate or server capacity) and concerns regarding tractability (note the transient analytical solution to even a basic *M*| *E*_*k*_| *1* queue is not without considerable complexity [26] – the queueing network considered here is of much greater sophistication).

The solution was to adopt a versatile open-source discrete event simulation tool that had recently been developed within the authors’ organisations (freely available at [27]). This provided the necessary functionality to accommodate all of the various modelling requirements articulated in Section 2.1. The tool implements the ‘three phase’ method to stochastic simulation [28], whereby discrete events are generated according to a schedule in which the next unconditional event is executed alongside any associated conditional events. An unconditional event represents either the arrival of a vaccinee at the site or an activity completion. If the former, then the generated conditional event is to enter service if capacity is available or to join a queue if otherwise. If the latter, then the conditional event generated is to exit service and present at the next service point or exit the site if the completed service represents the final activity on the vaccination pathway. The schedule is updated at each iteration and events continue until the end of the operating period is reached. In accounting for realistic conditions at the beginning of the operating period, each simulation starts empty and with no warm-up period.

Full results were obtained by performing multiple replications of the simulation, each with a different random number seed used to generate the timing of vaccinee arrivals and the service times at each activity. The former was sampled from a Poisson distribution (assuming independence of one arrival to another) and the latter from the most appropriate distribution for that activity. This was determined from fitting, by maximum likelihood estimation, the exponential, normal, lognormal, gamma and Weibull distributions to collected sample data for each activity, with selection by Akaike Information Criterion [29]. In generating results, 1500 replications were performed for each scenario as, following experimentation, this number was found to be sufficient in reducing simulation error on mean outcome measures to within 1% (an amount considered negligible for the intended use and given other sources of uncertainty).

## 3. Application

### 3.1 The initial setup of Vaccination Centre A

In December 2020, following news of the PZ vaccine success, efforts were being made to prepare the site ahead of planned public opening in early January 2021. Integral to this was a major live exercise conducted at the site (only the second of its kind to be run nationally) with support from the military and a host of volunteers who played out the vaccination process from start to finish. While this did not involve actual members of the public nor actual vaccinations being administered, the cast were provided with various scripts instructing them to act out a range of potential vaccinees and scenarios. Additionally, to promote realistic operating conditions, the site was organised according to the planned layout, which followed the ‘separate’ design (Figure 1). When opened to the public and running at full capacity, it was envisaged that there would be six registration assistants, 12 clinical assessors, and six vaccinators – which would be arranged in three 2-vaccinator ‘pods’. Post-vaccination observation space (when administering the PZ vaccine) would be set at 64. There was a finite queueing capacity of six and 15 ahead of *Clinical Assessment* and *Vaccination* respectively. At the time, national guidance was that a single 2-vaccinator pod could maintain a throughput of 520 vaccinees per 12-hour operating period – thus a total daily throughput of 1560 for the three-pod setup. The modelling objective was to assess, using data collected from the live exercise, the optimality of this figure and of the proposed resource configuration.

Various statistical distributions were fitted to sample service time data for *Registration* (n=20), *Clinical Assessment* (n=53) and *Vaccination* (n=24) through the approach described in Section 2.2 (noting that *Observation* service time was fixed at 15 minutes, as per clinical guidance for the PZ vaccine [30]). The best-fitting distributions were found to be Weibull, lognormal and lognormal respectively. Satisfactory goodness of fit was supported by the empirical data being contained within bootstrapped confidence limits (Figure 2). Parameter estimates and AIC for all considered distributions can be found in Supplementary Material A.

**Figure 2.**
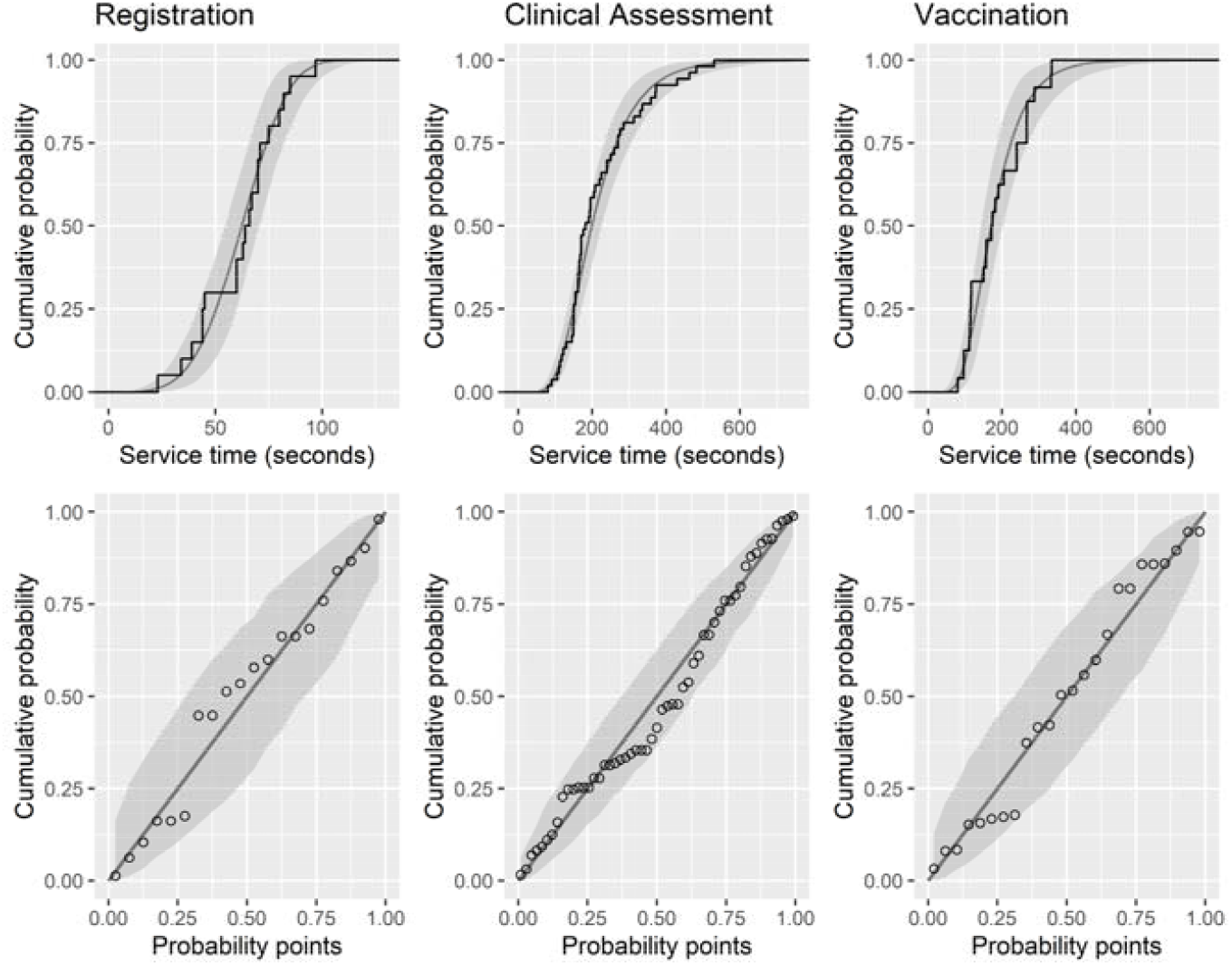
Cumulative distribution function and probability-probability plot showing the goodness of fit of the fitted distributions to empirical service time data for *Registration* (n=20), *Clinical Assessment* (n=53) and *Vaccination* (n=24), obtained from a live exercise ahead of the public opening of Vaccination Centre A.

Simulation results suggested that the envisaged configuration is unviable, with an increasingly large queue developing at *Vaccination* (Table 1, Baseline). This finding can actually be derived without modelling – 1560 arrivals simply cannot be accommodated with a pathway containing an activity whose maximum 12-hour throughput is only 1386 (i.e. from six vaccinators with a 187 second mean service time). Assuming vaccination time cannot be decreased, the solution was either to increase capacity or reduce arrivals. In order to safely accommodate the various peaks and troughs in arrivals and service time, the number of bookings should be sufficiently less than maximum throughput [31]. Lowering the 12-hour arrival rate by 10% from 1386 to 1247 results in performance within operational limits (Table 1, Scenario 1). It would, however, be prudent to increase the *Vaccination* waiting space (from 15) in order to absorb any potential ‘shocks’ relating to periods of elevated demand or staff shortages. Given spatial constraints of the site, this can be achieved by shifting the vaccination space into a reduced-capacity *Observation* space (noting that *Observation* capacity can be safely reduced since it is considerably under-utilised). *Registration* and *Clinical Assessment* were also under-utilised, implying uneconomic use of available resource. Modelling a one-sixth capacity reduction (i.e. to five and 10 posts respectively) is not shown to have an adverse performance impact (Table 1, Scenario 2).

**Table 1.** Simulated results for the number of individuals in service and in queue under the Baseline scenario and hypothetical Scenarios 1 (arrivals 10% less than maximum throughput) and 2 (arrivals 10% less than maximum throughput and one-sixth reductions to *Registration* and *Clinical Assessment* capacity). Values are steady-state results, achieved within the first hour of the 12-hour operating period, unless otherwise stated.

|  |  | Registration | Clinical Assessment | Vaccination | Observation |
| --- | --- | --- | --- | --- | --- |
| Mean number<br>(95% CI) of<br>vaccinees in<br>service | Baseline | 2.2 (0.0 – 5.6) | 7.8 (3.0 – 12.0) | 6.0 (6.0 – 6.0)* | 29.6 (24.7 – 34.4) |
|  | Scenario 1 | 1.8 (0.0 – 4.9) | 6.2 (2.0 – 11.6) | 5.1 (1.7 – 6.0) | 25.4 (16.8 – 32.3) |
|  | Scenario 2 | 1.8 (0.0 – 4.9) | 6.2 (2.0 – 10.0) | 5.1 (1.8 – 6.0) | 25.4 (16.9 – 32.4) |
| Mean number<br>(95% CI) of<br>vaccinees in<br>queue | Baseline | 0.0 (0.0 – 0.0) | 0.2 (0.0 – 2.2) | 108.0 (34.9 – 179.0)* | 0.0 (0.0 – 0.0) |
|  | Scenario 1 | 0.0 (0.0 – 0.0) | 0.0 (0.0–0.0) | 2.1 (0.0 – 11.6) | 0.0 (0.0 – 0.0) |
|  | Scenario 2 | 0.0 (0.0 – 0.0) | 0.1 (0.0–1.9) | 2.1 (0.0 – 11.1) | 0.0 (0.0 – 0.0) |

\* Values at end of 12-hour operating period. Behaviour did not stabilise during operating period.

Vaccination Centre A went live on 11 January 2021 with one vaccination pod open, increasing to two and then three in subsequent weeks. The number of individuals booked each day for vaccination, using the AZ vaccine, was based upon the afore-mentioned 1247 figure – that is, 416 bookings for each operational vaccination pod per day. Modelling also influenced the decision to increase the *Vaccination* queue capacity through a reduced *Observation* space. It was not possible to meaningfully validate site performance since real-life operation involved the periodic shutting down and reopening of various service channels. This meant that activity-level occupancy and queue size would fluctuate in ways not fully appreciated by the model. That said, clinical leads were supportive of the 416 figure in their assessment of achieving a good balance between high throughput and site safety. As such, it was used to set daily booking numbers throughout the opening month into mid-February 2021.

### 3.2 Transition to a ‘combined’ design at Vaccination Centre A

Data was collected from the site on 25 and 26 January 2021 with the objectives of reappraising throughput and resource configuration and assessing the potential value of a ‘combined’ design. While, at the time, the majority of vaccinees were routed through the ‘separate’ *Clinical Assessment* and *Vaccination* activities, some were selected to pass directly from *Registration* to one of two ‘combined’ service points (where they would be clinically assessed and vaccinated in one go). The site was trialling this option because of envisaged efficiency-savings relating to a smaller number of queues and eliminating duplication of certain questions and greetings exchanged with the vaccinee.

Arrival data was collected in order to validate the Poisson assumption. For arrivals to be Poisson-distributed then inter-arrival times must be exponentially-distributed. Adequate goodness of fit was supported by the empirical data (n=60) being contained within bootstrapped confidence limits (Figure 3). For service times, the lognormal distribution was selected as the best-fitting distribution across all activities (Table 2, Figure 4). Parameter estimates and AIC for all considered distributions can be found within Supplementary Material B.

**Table 2.**
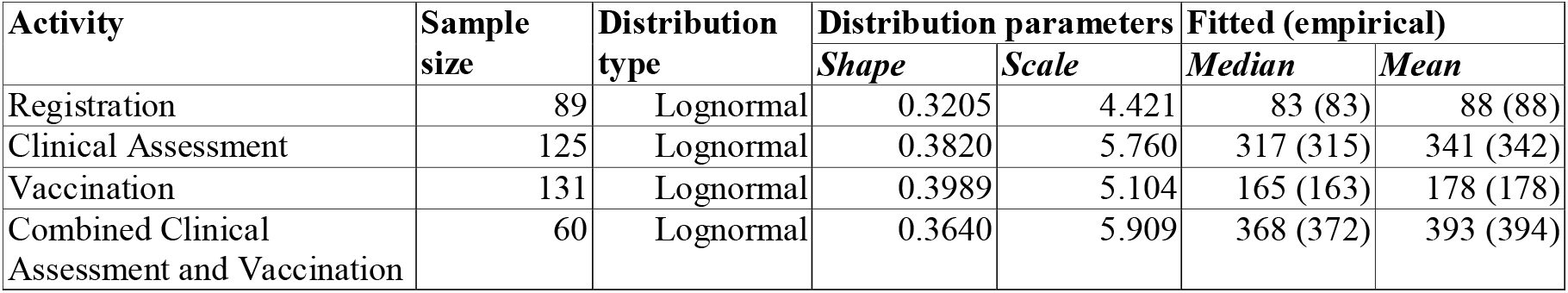
Distribution fits to empirical service time data obtained on 25 and 26 January 2021 at Vaccination Centre A.

**Figure 3.**
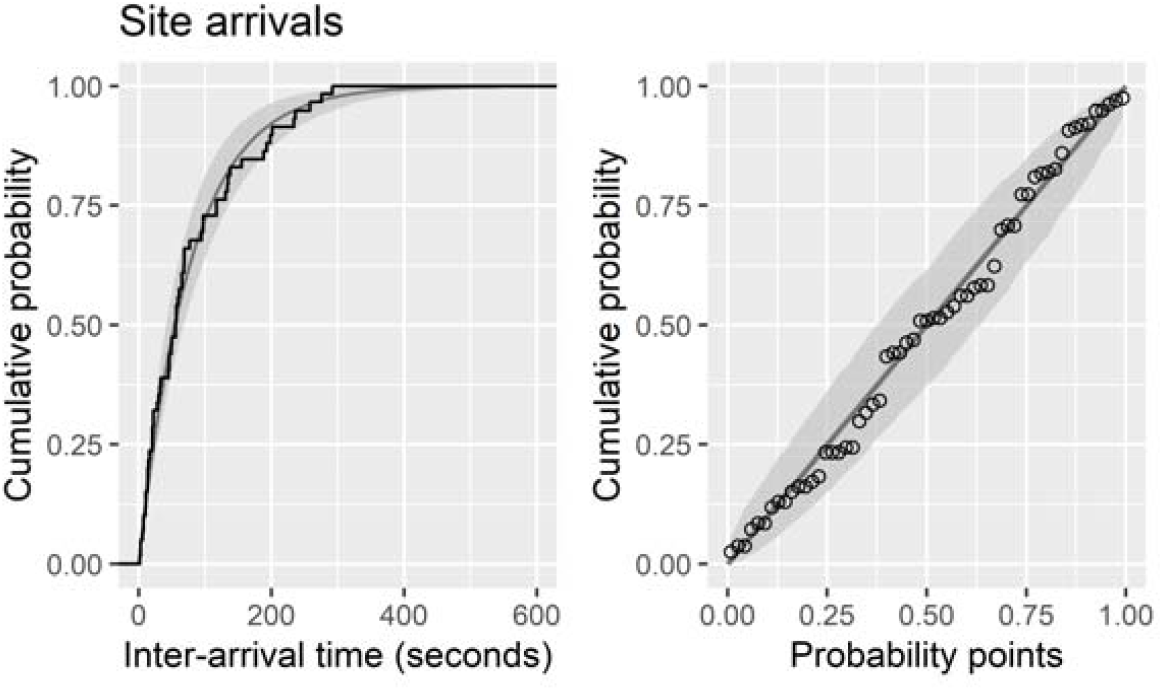
Cumulative distribution function and probability-probability plot showing the goodness of fit of the exponential distribution to empirical site inter-arrival time data (n=60), obtained on 25 January 2021 at Vaccination Centre A.

**Figure 4.**
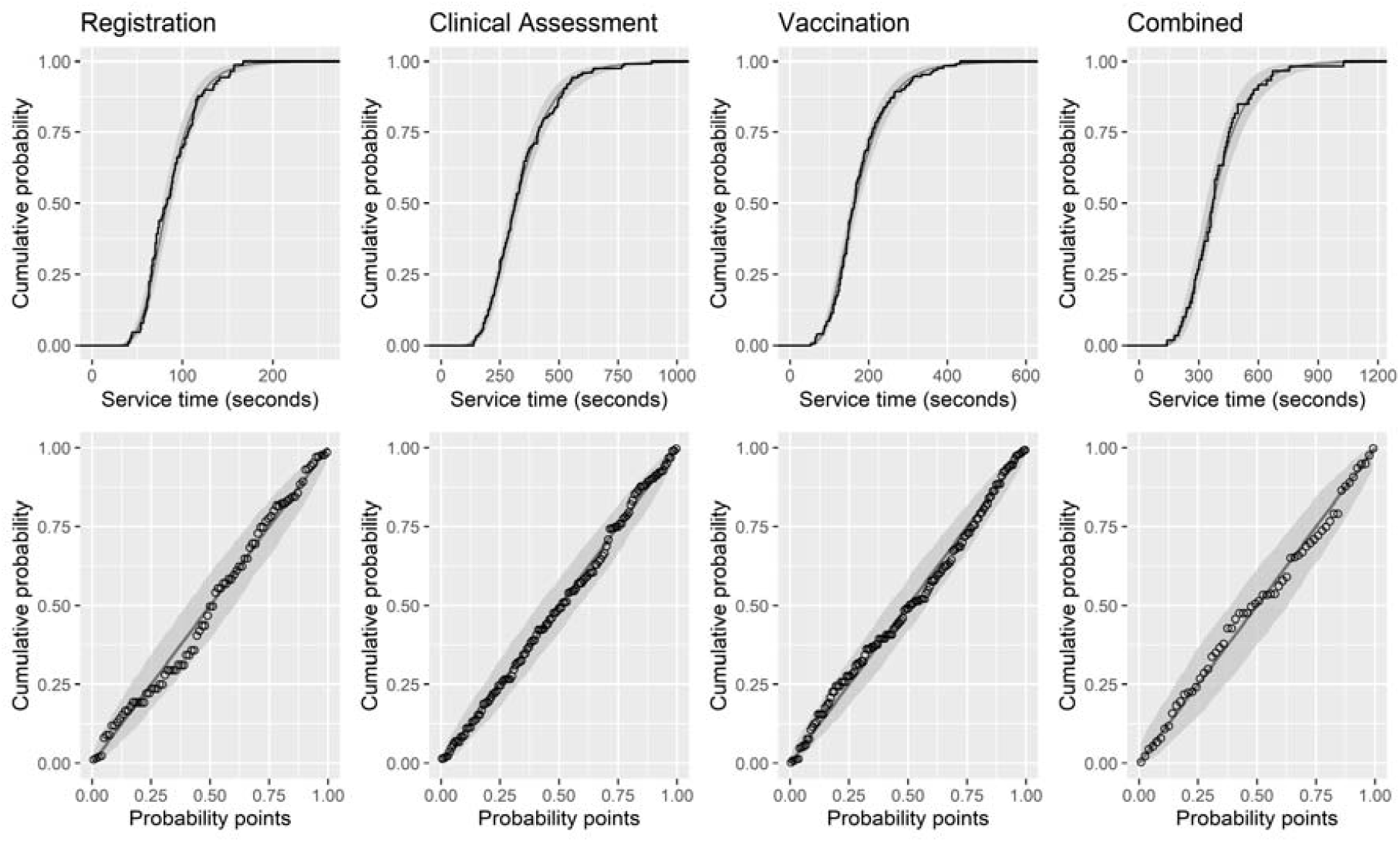
Cumulative distribution function and probability-probability plot showing the goodness of fit of the fitted distributions to empirical service time data for *Registration* (n=89), *Clinical Assessment* (n=125), *Vaccination* (n=131) and *Combined* (n=60), obtained on 25 and 26 January 2021 at Vaccination Centre A.

First, simulation results were obtained for the ‘separate’ configuration in reappraising the original throughput and capacity assessments as calculated using live exercise data (Section 3.1). As before, the number of 2-vaccinator pods was limited to three. Physical site constraints also translated into limited queueing capacity in the model, such that upper bounds on queue size for *Clinical Assessment* and *Vaccination* were approximately 10 and 20 respectively, and no more than 30 combined. Given the expected variability in queue size over time in the real-life system, it was acknowledged that these may, at certain times, be exceeded in the model. The tolerance was set such that the estimated upper 95% confidence band on queue size should not breach these limits during the simulated operating period. For the modelling, throughput was maximised under this condition. This yielded an estimated safe maximum throughput of 1330 arrivals per 12-hour operating period, with respective capacities of six and 13 for *Registration* and *Clinical Assessment* (Figure 5). This value is 7% greater than the original estimate, due to an equivalent reduction in (mean) vaccination service time.

**Figure 5.**
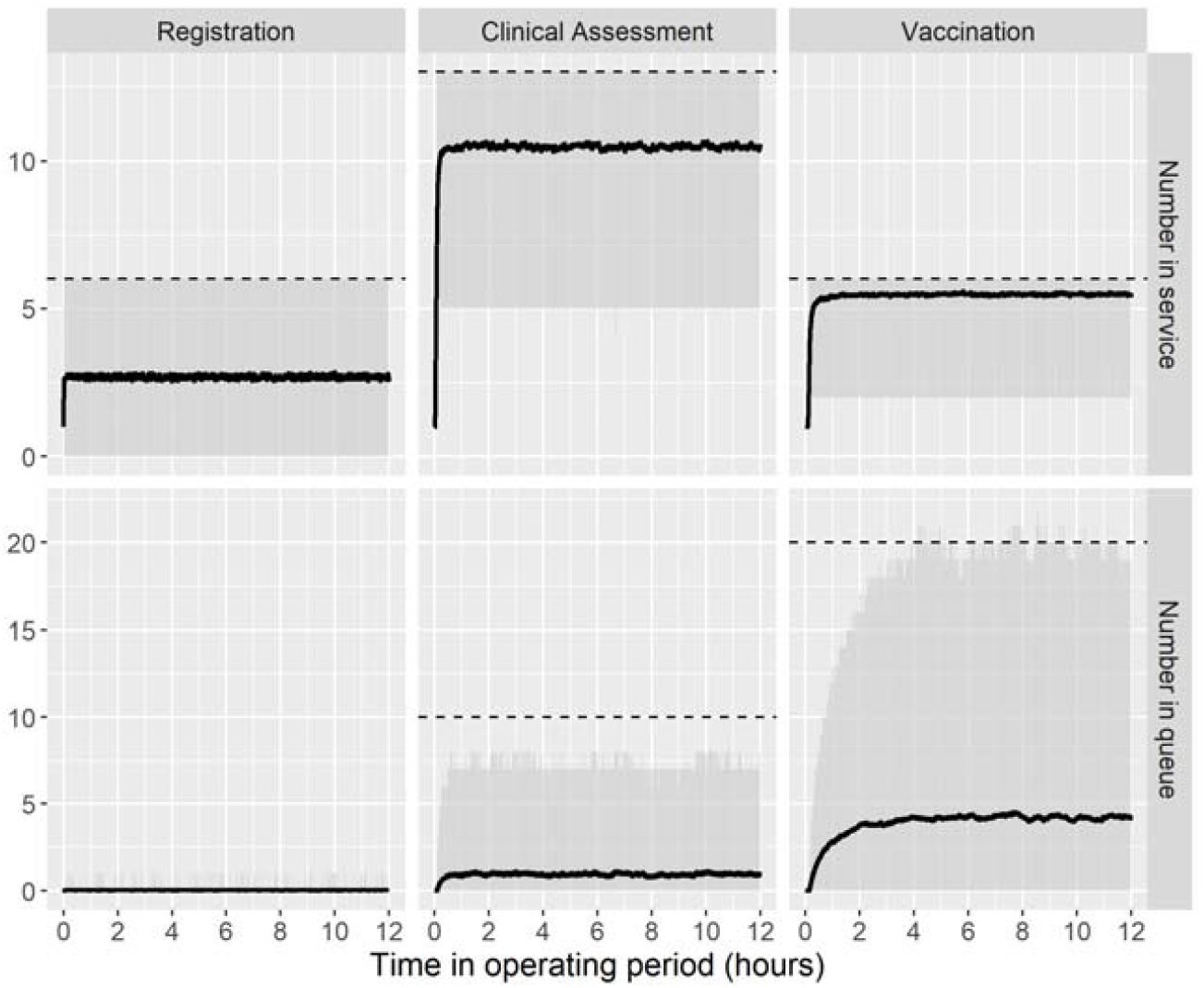
Simulated results for the number of individuals in service and in queue at Vaccination Centre A operating under the ‘separate’ design and with an optimal throughput of 1330 arrivals per 12-hour operating period. Dashed lines represent the activity-level service point and queue capacities.

Simulations results were next obtained for the ‘combined’ configuration, in order to assess its potential value. In order to ensure a fair comparison, the same limit (30) on the upper bound of queue size was used and human resources were constrained to the extent of those available under the above simulated ‘separate’ configuration (this meant respective capacities of six and 13 for *Registration* and *Combined*). Modelling indicated a throughput of 1340 could be achieved under such configuration (see Supplementary Material C for full simulation outputs). As well as achieving a (slightly) higher throughput (c.f. 1330), benefits would also extend to a reduced need for queueing with the vaccinee queueing a maximum of two, instead of three, times. This was important given that many of those being vaccinated were elderly and of reduced mobility (i.e. getting up and down multiple times and walking between different queues was undesirable). Two weeks following presentation of the analysis (2 February 2021), a decision was made to transition the site to a fully ‘combined’ configuration.

### 3.3 Performance of Vaccination Centre A vs Vaccination Centre B

Data was collected from Vaccination Centre B on 7 February 2021. Vaccination Centre B was already up-and-running at the time of involvement, operating under a fully ‘combined’ design and, on the day of the visit, using the AZ vaccine. The data indicated significantly reduced service times when compared to Vaccination Centre A (Table 2), with a mean duration of 47 (c.f. 88) seconds at *Registration* and 201 (c.f. 394) seconds at *Combined*. While the determinants of the lower times were clearly of interest, the immediate question from a modelling perspective was what degree of scope this offered for throughput improvements at Vaccination Centre A.

The ‘combined’ configuration of Section 3.2 was modelled as before but with the activity-level service time distributions replaced by those fitted to data from Vaccination Centre B (Supplementary Material D). With *Registration* and *Combined* service point capacities at six and 13 respectively, as before, this yielded a theoretical maximum throughput of 2620. This is 1280 (96%) higher than the equivalent 1340 figure, representing the achievable maximum at Vaccination Centre A at the time of the analysis. Results were relayed to management at Vaccination Centre A in order to provide an assessment of potential throughput gain were service times brought in line with those shown to be possible elsewhere. At the time of writing, opportunities to reduce service time were being considered. An option to reduce vaccination time may relate to the observation that much time was taken by vaccinees in removing multiple layers of (winter) clothing. Possible mitigations may include improved heating at the site, use of a cloakroom, or better signage indicating the need to remove excess clothing ahead of vaccination.

### 3.4 The impact of disruptions at Vaccination Centre B

Management had indicated the potential value in understanding the resilience of the vaccination pathway to unforeseen short-term disruptions, or ‘shocks’, relating to arrivals and site capacity. This would be useful information in determining the nature, magnitude and timing of any mitigation that may be required. While it was, given limited operational experience, not possible to define shocks in an empirically rigorous manner, two scenarios were discussed with operational managers and considered to be of reasonable plausibility. These were simulated against a baseline optimised on conditions observed at Vaccination Centre B on the day the data was collected, namely respective *Registration* and *Combined* capacities of five and 16 and a dedicated maximum *Combined* queueing capacity of 35. The first scenario involved a perturbation to the arrival rate such that half of those scheduled to arrive within the second hour of operation arrived within the third. This was intended to represent the possible disruption to site access caused by adverse travel conditions (such as a road traffic accident) occurring within the early morning rush hour period. The second scenario involved a perturbation to the *Combined* capacity such that one of the 16 service channels was unavailable for the first three hours of the operating period. This may represent the situation where a member of staff was absent with illness, until the time at which a temporary replacement can be found. The objective was to assess the responsiveness of operational performance, in terms of service point occupancy and queue size, to these considered shocks.

Simulated results for the first scenario are presented in Figure 6. These suggest that if just half of the bookings scheduled for one hour arrive in the next then there would be significant congestion at the site, with queues at *Registration* and *Combined* exceeding their typical safe limits (12 and 35 respectively). If this occurs early in the operating period, as modelled, then pressures, while subsiding, would persist for the remainder of the day. Simulated results for the second scenario are presented in Figure 7. This shows that, were one of the 16 *Combined* service channels not able to open until the fourth operating hour, then the upper 95% confidence level on the numbers queueing for *Combined* would exceed by two-fold the typical safe limit (35).

**Figure 6.**
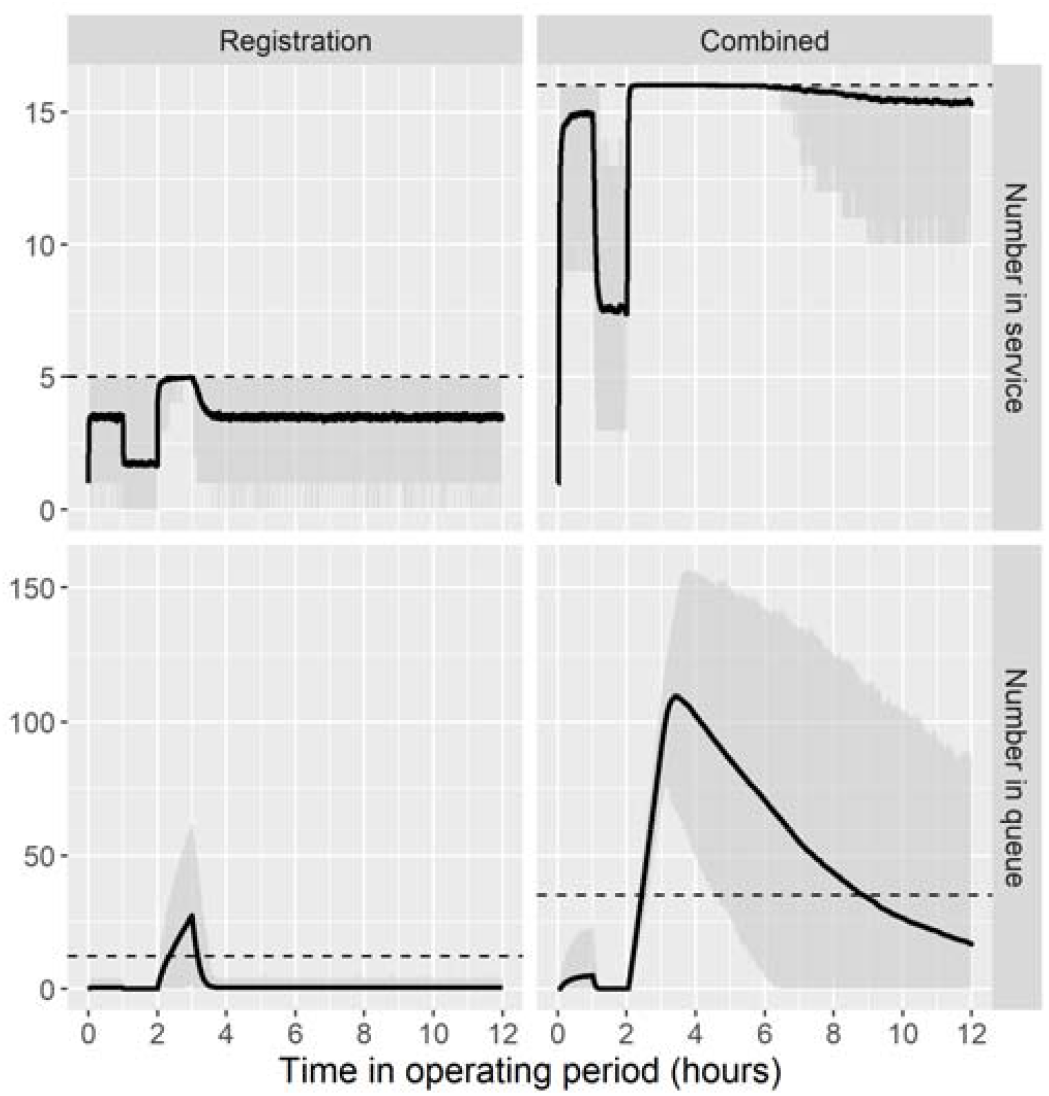
Simulated results for the number of individuals in service and in queue at Vaccination Centre B, with an adjusted arrival rate such that half of arrivals booked for the second operating hour arrive in the third.

**Figure 7.**
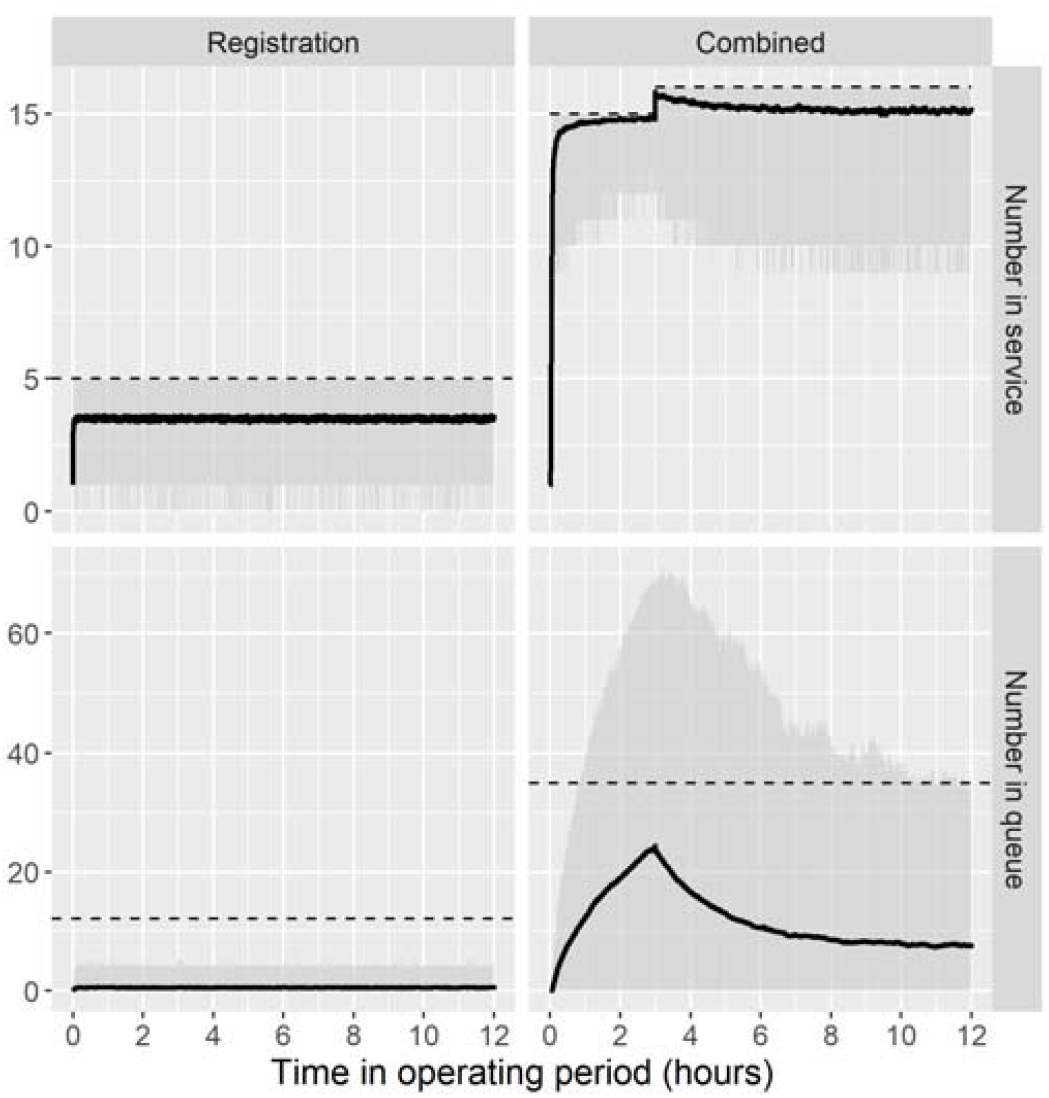
Simulated results for the number of individuals in service and in queue at Vaccination Centre B, with *Combined* capacity reduced by one service channel for the first three operating hours.

Consideration of both scenarios illustrates the potential perils of running a vaccination pathway optimised upon typical operating conditions. A possible solution could be to set daily bookings at a lower level, in order to provide sufficient ‘slack’ to safely absorb any such shocks when they do arise. An alternative solution is to ensure sufficient physical space is set aside for additional ‘overflow’ queueing capacity for use as and when required. Understanding the desirability of these mitigations would stem from a more detailed understanding and appraisal of management risk appetite, focusing on the extent to which lesser aggregate throughput is justifiable by reductions to the likelihood of significant operational disruptions.

## 4. Discussion

Through a multisite application, this study has shown that computer modelling can produce timely analysis of the appropriate sophistication to inform management considerations and operational decision making at COVID-19 vaccination centres. The analysis, performed at the very start of the UK’s mass vaccination effort (December 2020 to February 2021), has illustrated the capability of simulation to influence both the initial configuration of vaccination centres and their ongoing operation. One example is the setting of daily bookings at Vaccination Centre A. Adverse consequences were averted through use of the modelled 416 figure (for bookings per 2-vaccinator ‘pod’ per day). Had bookings been set 25% higher, at the originally proposed 520 figure, then capacity would likely have been overwhelmed, resulting in significant queueing at the site (Section 3.1). With many of the bookings initially allocated to the elderly and with much media interest, this could have generated negative publicity in the local and national press as well as compromising public confidence. Other examples in this paper evidence just some of the ways in which modelling and simulation can improve vaccination centre operations.

As with any modelling study, the various assumptions and simplifications can contribute to a number of limitations. With no reneging or balking or priority-based service order, the vaccination pathway actually represents a straightforward queueing network. Given also the evidence that arrivals are random (Poisson distributed) and that activity-level service times are adequately represented by the considered 2-parameter distributions, the limitations of this study relate mainly to application as opposed to conceptual or methodological appropriateness. Specifically, the conditions surrounding data collection should be carefully considered. First, vaccinees were typically over 70, since vaccination priority in the UK was based primarily upon decreasing age [10]. Second, vaccinations took place over winter, when additional clothing was being worn. Third, data was collected within four to six weeks of each site opening, when staff would not yet be fully familiarised to their new roles. Therefore, in the months following this study it would reasonably be expected that service times would reduce, which, in turn, would allow greater throughput than that deduced here. However, it is important to note that these three factors are not wholly explanative of service time variation, given the substantial differences in *Combined* duration between Vaccination Centres A and B (Section 3.3). This would therefore suggest caution in seeking to extend the specific results of this study to other sites.

In the long term, experimentation across the many vaccination centres being set up in the UK and further afield may reveal a set of optimal service models for which operational parameters can be determined and made available ‘off the shelf’ for individual sites adopting such configurations. Unless and until such time, vaccination centres seeking to improve operational efficiency should consider bespoke modelling as a viable option. As evidenced here, a key model requirement is versatility, in terms of accommodating different statistical distributions or service points along the pathway. In addition to the open-source model used in this study [27], many other simulation packages possess such versatility and so could be readily used. Those already familiar to healthcare analysts, at least in the UK, include Simul8 [32] and JaamSim [33]. Additionally, these programs would be sufficiently versatile in modelling 24-hours-a-day site operation, as has been suggested by the UK Government [34].

From an applied research perspective, given the paucity of the current literature and the possibility of mass vaccination becoming an annual exercise [35], there are many opportunities for future work in this problem area. Investigators may wish to assess whether the lognormal distribution continues to provide the best fit to service times in other settings and with other age groups. The extent to which combining *Clinical Assessment* and *Vaccination* improves efficiency through ‘economies of scale’ in pooling queueing and service time variance could also be investigated. While service point occupancy has been modelled in this study, further work could assess the impact on staff productivity – for instance, would the four hour period of 100% occupancy for the *Combined* service point (Figure 6) negatively affect the service rate, thus decreasing the pace at which the queue can be worked through? Researchers may also wish to consider the effect of vaccinee age on the tolerances that can be set for queuing. Provided physical space permits, a higher throughput could legitimately be achieved through allowing longer queues if those queueing were of greater resilience to withstand the additional waits. In this respect, investigators may seek to include some explicit measure of waiting time within any further analysis (noting that this is only equivalent to queue size for a given throughput [36]). Finally, future efforts could investigate the magnitude and effect of ‘no shows’ – with predictive analytics enabling the use of targeted reminders to those most at risk, and airline-style overbooking used to ensure that effective daily arrivals match as closely as possible the intended throughput.

## Data Availability

Data is available from the permanently hosted repository at:

https://github.com/nhs-bnssg-analytics/vacc-centre-data

## Acknowledgements

The authors acknowledge the contributions of Hannah Bailey, Karen Evans, Lucy Harries, Elizabeth Luckett, Anne Morris, Richard Rees, Mark Sanger, Trevor Shippey, Alex Thompson and Hayley Ware. The authors are also grateful to The Health Foundation (Evidence into Practice award) for funding this study.

## Supplementary Material A

Parameter estimates and model performance fitted to sample data for activity-level service times obtained from a live exercise conducted at Vaccination Centre A in advance of public opening. Values in seconds and to four significant figures.

| Activity | Fitted distribution | Parameter estimate |  | Akaike Information Criterion |
| --- | --- | --- | --- | --- |
|  |  | *Shape* | *Scale* |  |
| Registration<br>(n = 20) | Exponential | - | 0.01614 | 207.1 |
|  | Normal | 18.28 | 61.95 | 177.0 |
|  | Lognormal | 0.3450 | 4.073 | 181.1 |
|  | Gamma | 9.618 | 0.1553 | 179.1 |
|  | Weibull | 3.915 | 68.53 | 176.4 |
| Clinical Assessment<br>(n = 53) | Exponential | - | 0.004554 | 679.5 |
|  | Normal | 102.4 | 219.6 | 645.1 |
|  | Lognormal | 0.4285 | 5.296 | 626.0 |
|  | Gamma | 5.400 | 0.0246 | 629.7 |
|  | Weibull | 2.281 | 248.9 | 637.4 |
| Vaccination<br>(n = 24) | Exponential | - | 0.005368 | 300.9 |
|  | Normal | 75.28 | 186.3 | 279.5 |
|  | Lognormal | 0.4136 | 5.144 | 276.6 |
|  | Gamma | 6.131 | 0.03291 | 276.8 |
|  | Weibull | 2.694 | 210.3 | 277.8 |

## Supplementary Material B

Parameter estimates and model performance fitted to sample data for activity-level service times obtained from Vaccination Centre A on 25 and 26 January 2021. Values in seconds and to four significant figures.

| Activity | Fitted distribution | Parameter estimate |  | Akaike Information Criterion |
| --- | --- | --- | --- | --- |
|  |  | *Shape* | *Scale* |  |
| Registration<br>(n = 89) | Exponential | - | 0.01142 | 976.1 |
|  | Normal | 28.69 | 87.55 | 854.0 |
|  | Lognormal | 0.3205 | 4.421 | 840.9 |
|  | Gamma | 9.858 | 0.1126 | 842.8 |
|  | Weibull | 3.213 | 97.72 | 853.5 |
| Clinical Assessment<br>(n = 125) | Exponential | - | 0.002927 | 1710 |
|  | Normal | 137.3 | 341.6 | 1589 |
|  | Lognormal | 0.3820 | 5.760 | 1558 |
|  | Gamma | 6.924 | 0.02027 | 1562 |
|  | Weibull | 2.599 | 384.9 | 1581 |
| Vaccination<br>(n = 131) | Exponential | - | 0.005611 | 1622 |
|  | Normal | 73.80 | 178.2 | 1503 |
|  | Lognormal | 0.3989 | 5.104 | 1472 |
|  | Gamma | 6.459 | 0.03624 | 1475 |
|  | Weibull | 2.528 | 201.1 | 1492 |
| Combined<br>(n = 60) | Exponential | - | 0.002540 | 839.1 |
|  | Normal | 151.8 | 393.7 | 777.0 |
|  | Lognormal | 0.3640 | 5.909 | 762.0 |
|  | Gamma | 7.635 | 0.01939 | 764.0 |
|  | Weibull | 2.652 | 442.3 | 774.3 |

## Supplementary Material C

Simulated results for the number of individuals in service and in queue at Vaccination Centre A, operating under the ‘combined’ design and with an optimal throughput of 1340 arrivals per 12-hour operating period. Dashed lines represent the activity-level service point and queue capacities.

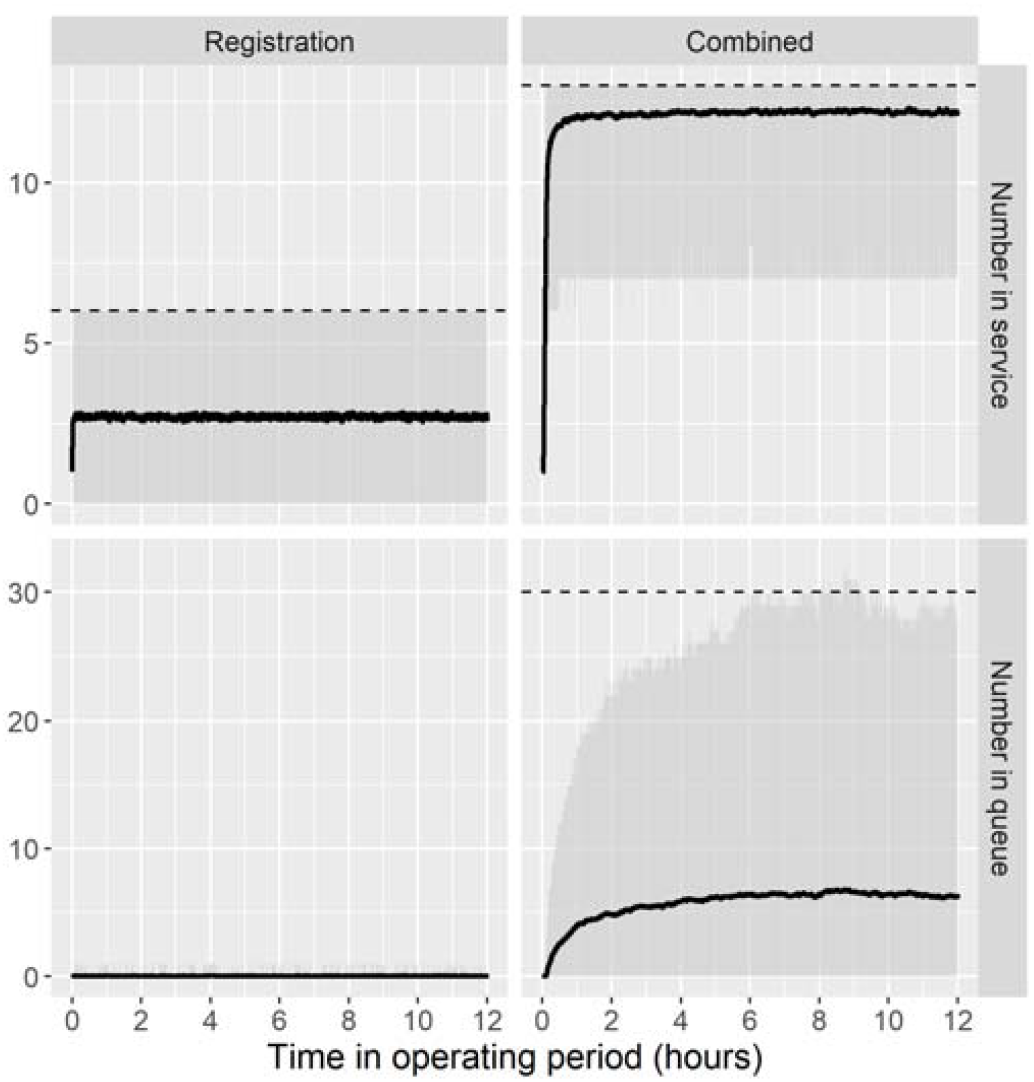

## Supplementary Material D

Parameter estimates and model performance fitted to sample data for activity-level service times obtained from Vaccination Centre B on 7 February 2021. Values in seconds and to four significant figures.

| Activity | Fitted distribution | Parameter estimate |  | Akaike Information Criterion |
| --- | --- | --- | --- | --- |
|  |  | Shape | Scale |  |
| Registration<br>(n = 70) | Exponential | - | 0.02141 | 680.1 |
|  | Normal | 16.44 | 46.70 | 599.1 |
|  | Lognormal | 0.2760 | 3.800 | 558.9 |
|  | Gamma | 11.63 | 0.2490 | 569.4 |
|  | Weibull | 2.684 | 52.10 | 599.8 |
| Combined<br>(n = 90) | Exponential | - | 0.004974 | 1137 |
|  | Normal | 84.97 | 201.0 | 1059 |
|  | Lognormal | 0.3877 | 5.225 | 1029 |
|  | Gamma | 6.524 | 0.03245 | 1036 |
|  | Weibull | 2.497 | 227.3 | 1050 |

Cumulative distribution function and probability-probability plot showing the goodness of fit of fitted lognormal distributions to empirical service time data obtained on 7 February 2021 at Vaccination Centre B.

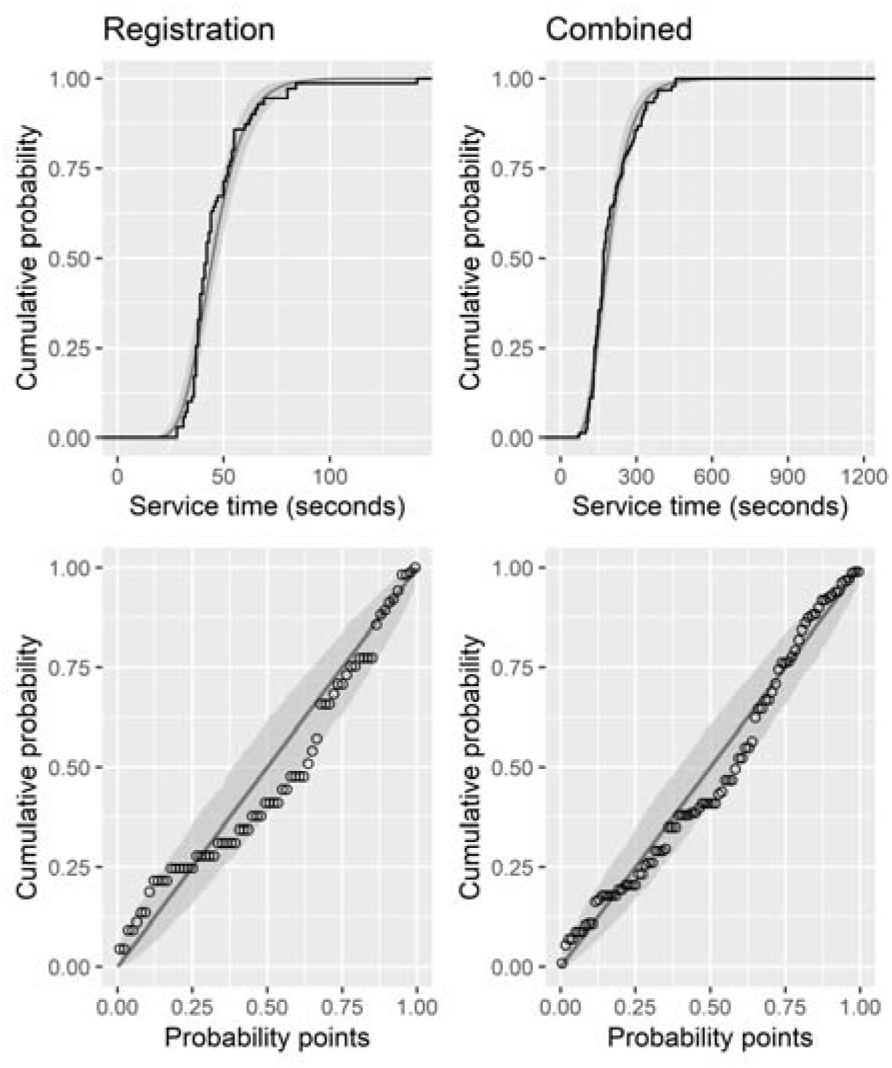

